# Designing a flow-controlled STA-MCA anastomosis based on the Hagen-Poiseuille law for preventing postoperative hyperperfusion in adult moyamoya disease

**DOI:** 10.1101/2022.06.04.22275816

**Authors:** Jianjian Zhang, Miki Fujimura, Tsz Yeung Lau, Jincao Chen

**Author notes:** **Corresponding Author contact information:** Jincao Chen, MD, PhD and Jianjian Zhang MD, PhD. Department of Neurosurgery, Zhongnan Hospital of Wuhan University, Donghu Road 169#, Wuhan, 430071, China; and.

## Abstract

**OBJECTIVE:** Technical improvements for preventing postoperative symptomatic cerebral hyperperfusion (CHP) during superficial temporal artery-middle cerebral artery (STA-MCA) anastomosis for moyamoya disease (MMD) were seldom reported. The aim of this study was to investigate the significance of application of a novel flow-controlled concept which voluntarily reduces the hemodynamic difference between the donor and recipient arteries based on the Hagen-Poiseuille law when performing direct anastomoses of recipient parasylvian cortical arteries (PSCAs) with anterograde hemodynamic sources from the MCA (M-PSCAs) in adult MMD.

**METHODS:** Recently direct anastomoses of recipient M-PSCAs were performed on 89 symptomatic hemispheres in 82 adult MMD patients in our hospital. They were divided into the flow-controlled group and non-flow-controlled group. The patients’ basic characteristics and incidence of postoperative CHP were compared between the two groups. Risk factors for occurrence of postoperative CHP were analyzed.

**RESULTS:** The earlier 36 and later 53 anastomoses were respectively included in the non-flow-controlled group and flow-controlled group. The incidences of postoperative focal (22.6%) and symptomatic CHP (5.7%) in the flow-controlled group were significantly lower than those (focal, 52.8%; symptomatic, 25.0%) in the non flow-controlled group (P = 0.003 and 0.009, respectively). Multivariate analysis revealed that the flow-controlled concept used or not was significantly associated with the development of focal (P = 0.005) and symptomatic (P = 0.012) CHP.

**CONCLUSIONS:** The flow-controlled STA-MCA anastomosis can significantly decrease the incidence of postoperative CHP during direct anastomoses of recipient M-PSCAs in adult MMD.

## Background

Moyamoya disease (MMD) is a chronic, occlusive cerebrovascular disease of unknown etiology characterized by bilateral stenotic-occlusive changes at the terminal portion of the internal carotid artery and an abnormal vascular network at the base of the brain.^11^ Direct revascularizations, such as superficial temporal artery-middle cerebral artery (STA-MCA) anastomosis, play a major role in immediately augmenting blood flow to the oxygen-deprived brain in MMD patients.^14^ Conventionally, to improve ischemic conditions of the hypo-perfused brain, neurosurgeons preferred to maximally achieve the goal of “flow augmentation” during the bypass procedures. However, with recent advances in microsurgical techniques, postoperative symptomatic cerebral hyperperfusion (CHP) becomes a big issue during STA-MCA anastomoses for adult MMD patients, it could result in transient neurological deficits (TND) and even cerebral hemorrhage.^3,5,6,15,20^ It seems that there lacks effective surgical treatments to prevent the appearance of postoperative symptomatic CHP under the traditional STA-MCA bypass procedure.

In the present study, as a continuation of our previous research^23^, we put forward a new flow-controlled concept of STA-MCA anastomosis when performing direct anastomoses of parasylvian cortical arteries (PSCAs) with anterograde hemodynamic sources from the MCA (named M-PSCAs). Because this kind of direct anastomoses were demonstrated to have a high risk of postoperative CHP,^23^ this new concept suggested a “controlled”, rather than a “maximized” augmentation of blood flow through reducing the hemodynamic difference between the donor and recipient arteries based on the Hagen-Poiseuille equation during their surgical procedures. The aim of this study is to analyze whether application of this kind of flow-controlled anastomosis decreased the incidence of the postoperative symptomatic CHP or not in adult MMD.

## Methods

### Patients

We retrospectively collected data of adult patients with MMD who underwent STA-MCA anastomosis combined with encephaloduromyosynangiosis (EDMS) revascularization in our hospital from January 2020 to June 2021. The diagnosis for each patient was established by an 8-vessel (bilateral internal carotid arteries, external carotid arteries, common carotid arteries and vertebral arteries) digital subtraction angiography (DSA). All patients satisfied the diagnostic criteria of the Research Committee on Spontaneous Occlusion of the Circle of Willis of the Ministry of Health, Labor, and Welfare, Japan.^7,18^ The magnetic resonance imaging (MRI) and angiography (MRA) scans were performed before surgery and within 1 week after surgery in each patient to evaluate underlying structural brain abnormalities and confirm the patency of the anastomosis. The cerebral perfusion status was routinely assessed by 99mTc-ECD single-photon emission computed tomography (99mTc-ECD-SPECT) before and one day after surgery in all patients.

The inclusion criteria of patients in this study were strict as patients who were performed direct anastomoses of recipient PSCAs with their hemodynamic sources from the MCA (M-PSCAs). Patients with blood flow of the recipient PSCAs from the non-MCAs were excluded from the study. The method for analyzing hemodynamic sources of PSCAs was reported elsewhere.^23^ Additionally, patients with aneurysms were also excluded because different surgical approaches were used during their revascularization surgeries (Figure 1).

**Figure 1.**
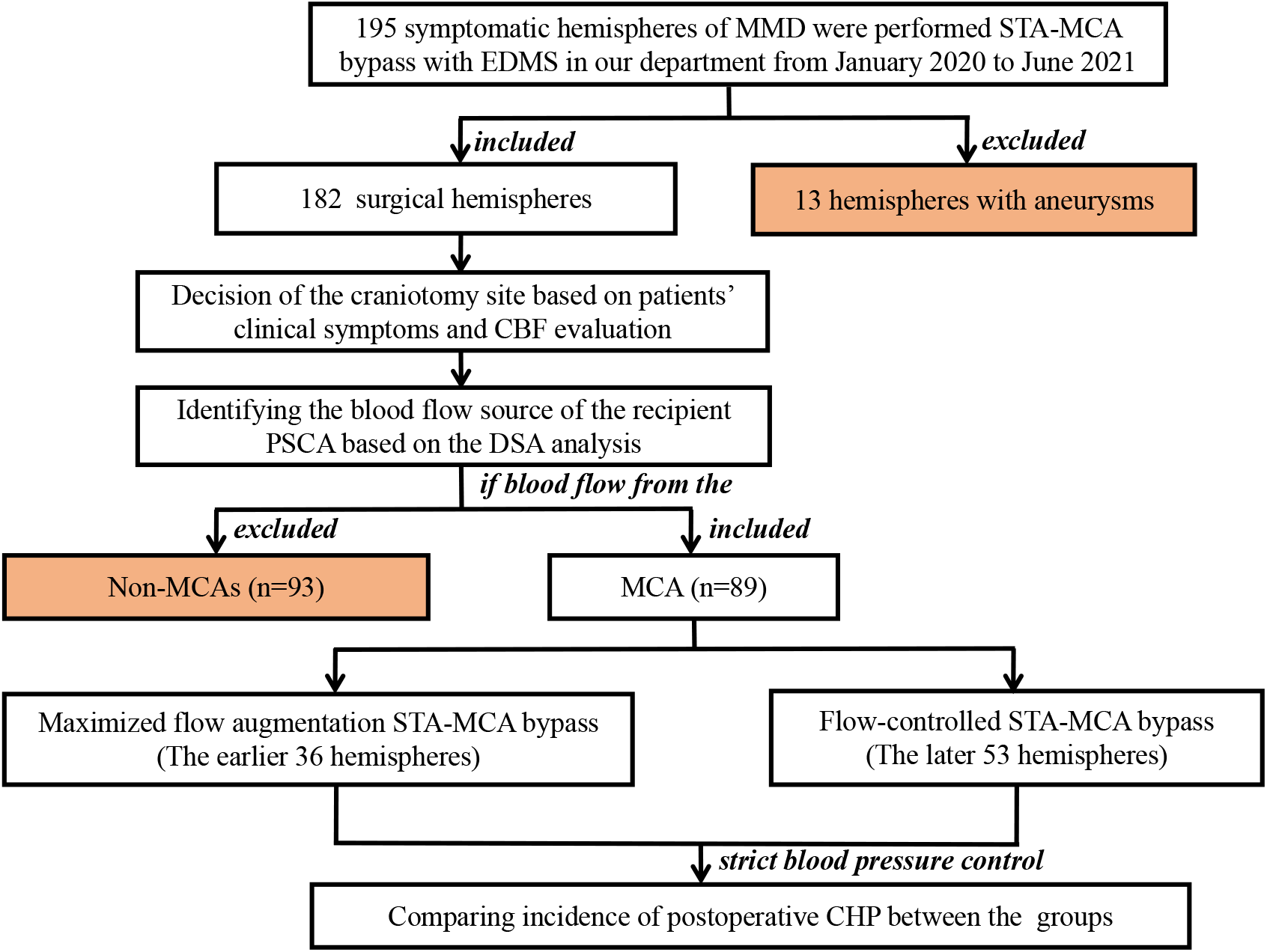
Patient inclusion flowchart. MMD, moyamoya disease; STA, superficial temporal artery; MCA, middle cerebral artery; EDMS, encephaloduromyosynangiosis; CBF, cerebral blood flow; PSCA, parasylvian cortical artery; 3D DSA-MR fusion imaging, three-dimensional (3D) digital subtraction angiography (DSA)-magnetic resonance (MR) fusion imaging; CHP, cerebral hyperperfusion.

This study protocol was approved by the Institutional Review Board at Zhongnan Hospital of Wuhan University (approval number: Kelun-2017005) and was in accordance with the Declaration of Helsinki revised in 1983. Written informed consent was obtained from all patients.

### Surgical Procedure and postoperative management

The indication for surgery included patients with symptomatic MMD (ischemic or hemorrhagic) with apparent hemodynamic compromise. The craniotomy was performed around the Sylvian fissure end to obtain more suitable recipient arteries on the cortical surface of the craniotomy site. The technical note of the traditional maximized flow augmentation STA-MCA anastomosis was already introduced by previous articles and books.^2,22,24^ The definition of focal (both asymptomatic and symptomatic) CHP and postoperative management for preventing CHP were as described elsewhere.^4,5,23^

### The flow-controlled STA-MCA anastomosis

The basic principle of the “flow-controlled” STA-MCA anastomosis was to reduce the hemodynamic difference between the donor and recipient arteries and ensure forward blood flow from the donor to the recipient at the same time. According to the famous Hagen-Poiseuille equation^8^: Q=π×r^4^×Δp/(8ηL), we designed the procedure of “flow-controlled” as follows:

1. Controlling the “L” in the equation: If there was a significant difference between the diameter of the frontal and parietal branches of the STA, the thinner one would be selected as the donor artery while ensuring its diameter > 1mm and the length of the final donor artery > 10 cm. Dissection of the STA branch with sufficient length could ensure enough small diameter of the stump of the STA branch suitable for the next controlling procedure.
2. Controlling the “Δp” in the equation: We kept the patency of the STA when dissecting it from the scalp and during the craniotomy. Before anastomosis, a flow 800 indocyanine green (ICG) video angiography was performed to observe the velocity difference between the STA and the cortical arteries. If there was a significant velocity difference between the M-PSCAs, we selected the artery with the closest blood flow velocity to the STA as the recipient vessel on the cerebral surface. However, it needed to be guaranteed that all the anastomoses were performed on M-PSCAs responsible for the hypoperfused brain.
3. Controlling the “r” in the equation: First, avoiding dilation of the STA when rinsing the lumen by heparin salt solution. Second, a micro-ruler was used to measure the diameters of STA and potential recipient M-PSCAs under microscope. Select one of the M-PSCAs with its diameter about or near to 1mm as the recipient artery.^9^ Third, designing the length of the arteriotomy less than 1.2-fold the diameter of the recipient artery.

### Statistical Analysis

A one-way ANOVA test was performed to see if there were any statistical differences of age and systolic blood pressure (SBP) between the groups. Categorical variables, such as sex, Suzuki stage, surgical side, initial onset type, focal CHP and symptomatic CHP were analyzed in contingency tables with Chi-square test. Multivariate statistical analysis of the factors related to symptomatic CHP was performed using a logistic regression model. All analyses were performed with IBM SPSS Statistics Desktop, version 24 (IBM Corp.). The results with values of *P*<0.05 were considered significant.

## Results

### Grouping and patient’s basic characteristics

Eighty-two adult patients with MMD (patients aged 19-65 years old, mean 47.03 years old) met the inclusion criteria and were retrospectively collected in this study. A total 89 symptomatic hemispheres of these patients were performed direct STA-MCA (M4) anastomosis combined indirect EDMS. Consequently, the earlier 36 hemispheres were included in the non-flow-controlled group while the later 53 hemispheres in the flow-controlled group. The basic characteristics of the hemispheres are summarized in Table 1.

**Table 1.** Comparison of basic characteristics and incidence of postoperative CHP between two groups.

|  | “Maximized flow augmentation” bypass<br>(n=36) | “Flow-controlled ” bypass<br>(n=53) | *P* Value |
| --- | --- | --- | --- |
| Age, y | 46.75 ± 11.30 | 47.23 ± 9.32 | 0.829 |
| Male sex (%) | 20 (55.6) | 20 (37.7) | 0.097 |
| Hemorrhagic onset (%) | 11 (30.6) | 21 (39.6) | 0.382 |
| Surgical side (lt, %) | 18 (50.0) | 27 (50.9) | 0.930 |
| SBP before surgery | 126.81 ± 15.03 | 128.68 ± 14.70 | 0.560 |
| SBP one day after surgery | 121.33 ± 11.72 | 123.17 ± 8.98 | 0.405 |
| Suzuki Stage (%) |  |  | 0.148 |
| 2 | 4 (11.1) | 9 (17.0) |  |
| 3 | 17 (47.2) | 33 (62.3) |  |
| 4 | 14 (38.9) | 11 (20.7) |  |
| 5 | 1 (2.8) | 0 (0) |  |
| Focal CHP | 19 (52.8) | 12 (22.6) | 0.003 |
| Symptomatic CHP | 9 (25.0) | 3 (5.7) | 0.009 |

CHP = Cerebral hyperperfusion. SBP = systolic blood pressure.

### Comparison of incidence of postoperative CHP between both groups

As summarized in Table 1, there were no significant differences of age, sex, initial onset type, surgical side, SBP before surgery, SBP one day after surgery and Suzuki stage between the flow-controlled group and non-flow-controlled group (P all>0.05). In the flow-controlled group, the incidence of postoperative focal CHP detected by SPECT was only 22.6% (12/53 hemispheres), which was significantly lower than that (52.8%, 19/36 hemispheres) in the non-flow-controlled group (*P* = 0.003). More importantly, the number of patients who developed symptomatic CHP in the flow-controlled group (3 patients, 5.7%) was significantly less than that (9 patients, 25.0%) in the non-flow-controlled group (P = 0.009). The representative image of the qualitative analysis of hemodynamic change from SPECT revealed significant intense increases in CBF at the sites of anastomosis (Figure 2A). Postoperative MRI/MRA showed no ischemic changes, and the thick high signal intensity of STA on the operated hemisphere was evident in all 89 hemispheres (Figure 2B). The 6 month follow up MRA showed graft occlusion occurred in 2 (2/53, 3.8%) hemispheres. Together, the rates of bypass graft patency during the operation and 6 month follow up were 100% and 96.2%, respectively.

**Figure 2.**
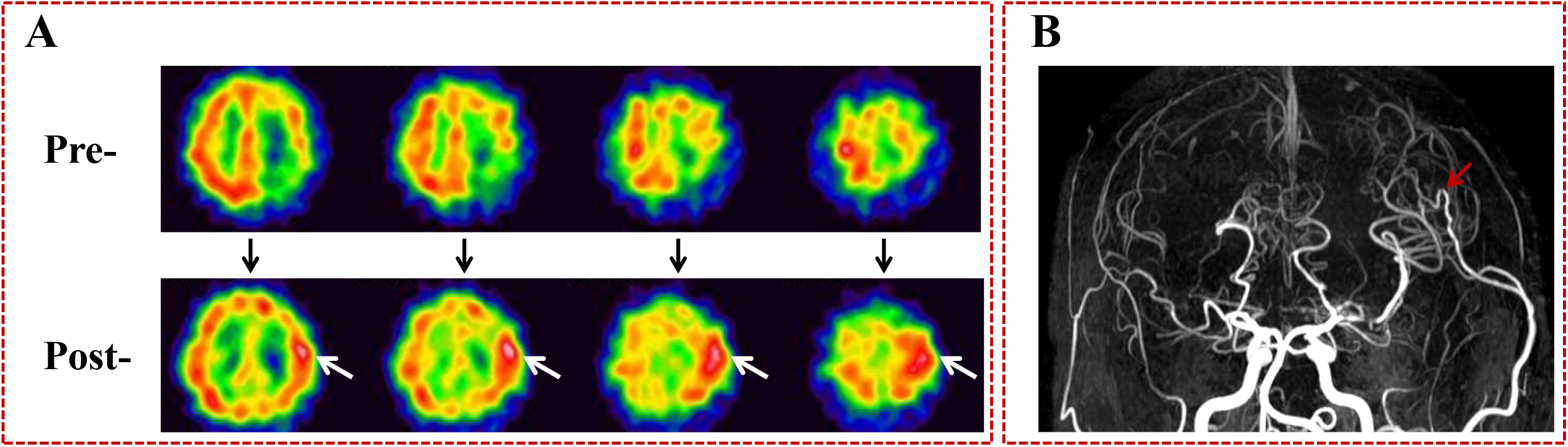
Representative images of focal cerebral hyperperfusion. **(A)** 99mTc-ECD single-photon emission computed tomography (99mTc-ECD-SPECT) one day after left STA-MCA anastomosis with EDMS. Focal increase in cerebral blood flow (CBF) was evident at the site of the anastomosis (white arrows in A). **(B)** MRA two days after operation, demonstrating high signal intensity of the STA branch (red arrow in B).

Among the 33 patients with 36 operated hemispheres of the non-flow-controlled group, 9 patients (9 hemispheres) occurred symptomatic CHP. The clinical presentations of symptomatic CHP included small cerebral hematoma (7ml) in 1 patient, moderate cerebral hematoma (20ml) in 1 patient, aphasia in 5 patients, seizure in 2 patients, and motor weakness in 7 patients. Most of the patients completely recovered, except that the patient with moderate cerebral hematoma developed permanent neurological deficits (Table 2).

**Table 2.** Clinical presentations and outcome of symptomatic CHP.

| No. | Age range | Sex | Surgical side | Suzuki's stage | Onset type | "Flow-controlled" | Clinical presentations of symptomatic CHP | Prognosis |
| --- | --- | --- | --- | --- | --- | --- | --- | --- |
| 1 | 20-25 | Male | Right | 3 | Ischemic | Yes | Dysarthria, seizure | Good |
| 2 | 46-50 | Female | Left | 4 | Ischemic | Yes | Motor aphasia | Good |
| 3 | 46-50 | Female | Left | 3 | Ischemic | Yes | Motor aphasia, weakness of the right limbs | Good |
| 4 | 50-55 | Male | Right | 4 | Ischemic | No | Seizure, weakness of the left limbs | Good |
| 5 | 15-20 | Female | Left | 3 | Hemorrhagic | No | Motor aphasia, weakness of the right limbs | Good |
| 6 | 50-55 | Male | Left | 5 | Ischemic | No | Moderate hematoma in the bypass site (20ml), motor aphasia and right hemiplegia | Disabled |
| 7 | 50-55 | Female | Right | 2 | Ischemic | No | Sensory aphasia, weakness of the right limbs | Good |
| 8 | 46-50 | Female | Left | 2 | Ischemic | No | Motor aphasia | Good |
| 9 | 46-50 | Male | Left | 4 | Hemorrhagic | No | Small hematoma near the bypass site (7ml) | Good |
| 10 | 51-55 | Female | Left | 4 | Ischemic | No | Numbness and weakness of the right limbs | Good |
| 11 | 46-50 | Female | Right | 3 | Ischemic | No | Seizure, weakness of the left limbs | Good |
| 12 | 56-60 | Female | Left | 3 | Ischemic | No | Motor aphasia, weakness of the right limbs | Good |

CHP = Cerebral hyperperfusion; ml = milliliter.

In comparison, among the 49 patients with 53 operated hemispheres of the flow-controlled group, only 3 patients (3 hemispheres) suffered from temporary neurological deterioration due to postoperative focal CHP (Table 2). Importantly, no intraoperative or postoperative cerebral hemorrhage occurred in this group.

The results of the multivariate analysis revealed that the flow-controlled procedure was significantly associated with the development of focal (P = 0.005) and symptomatic (P = 0.012) CHP. Moreover, other factors such as age, sex, surgical side, initial onset type, and Suzuki stage were not associated with both focal and symptomatic CHP (Table 3).

**Table 3.** Multivariate analysis of postoperative CHP after direct anastomoses of recipient PSCAs with anterograde hemodynamic sources from the MCA.

| Factors | Focal CHP |  | OR (95% CI) | P value | Symptomatic CHP |  | OR (95% CI) | P value |
| --- | --- | --- | --- | --- | --- | --- | --- | --- |
|  | Yes<br>(n=31) | No<br>(n=58) |  |  | Yes<br>(n=12) | No<br>(n=77) |  |  |
| Age, y | 47.13 ± 10.88 | 46.98 ± 9.77 | 0.997 (0.952-1.045) | 0.913 | 45.92 ± 12.03 | 47.21 ± 9.86 | 0.967 (0.908-1.029) | 0.286 |
| Male sex (%) | 12 (38.7%) | 28 (48.3%) | 2.340 (0.828-6.613) | 0.109 | 4 (33.3%) | 36 (46.8%) | 2.652 (0.594-11.849) | 0.202 |
| Suzuki stage (2/3/4/5) | 4/15/11/1 | 9/35/14/0 | 1.445 (0.696- 3.002) | 0.323 | 2/5/4/1 | 11/45/21/0 | 1.520 (0.558- 4.140) | 0.412 |
| Surgical side (lt, %) | 15 (48.4%) | 30 (51.7%) | 1.324 (0.508- 3.451) | 0.565 | 8 (66.7%) | 37 (48.1%) | 0.375 (0.086- 1.639) | 0.192 |
| Hemorrhagic onset (%) | 10 (32.3%) | 22 (37.9%) | 1.095 (0.402- 2.984) | 0.860 | 2 (16.7%) | 30 (39.0%) | 3.786 (0.636- 22.543) | 0.144 |
| Flow-controlled /maximized flow augmentation | 12/19 | 41/17 | 0.238 (0.088-0.644) | 0.005 | 3/9 | 50/27 | 0.140 (0.030-0.647) | 0.012 |

CHP = Cerebral hyperperfusion; PSCAs = parasylvian cortical arteries; MCA = middle cerebral artery; OR = Odd Ratio; CI = Confidence Interval.

### Illustrative Case

This case was a young man (36-40 years old) who presenting with recurrent weakness of the left limbs for 3 months and diagnosed with MMD by DSA. Although no cerebral infarct was observed from the MRI images, 99mTc-ECD-SPECT showed there was significantly decreased CBF in the right frontal and parietal lobes. The DSA demonstrated that blood flow of the PSCAs above the sylvian fissure totally came from the anterograde MCA. Then we decided to perform a flow-controlled anastomosis on a PSCA above the sylvian fissure of the right hemisphere (the detailed procedure was introduced in Figure 3). The temporary occlusion time was 16 minutes. Postoperative blood pressure was maintained in the normal range, and he did not present any neurological deficit perioperatively.

**Figure 3.**
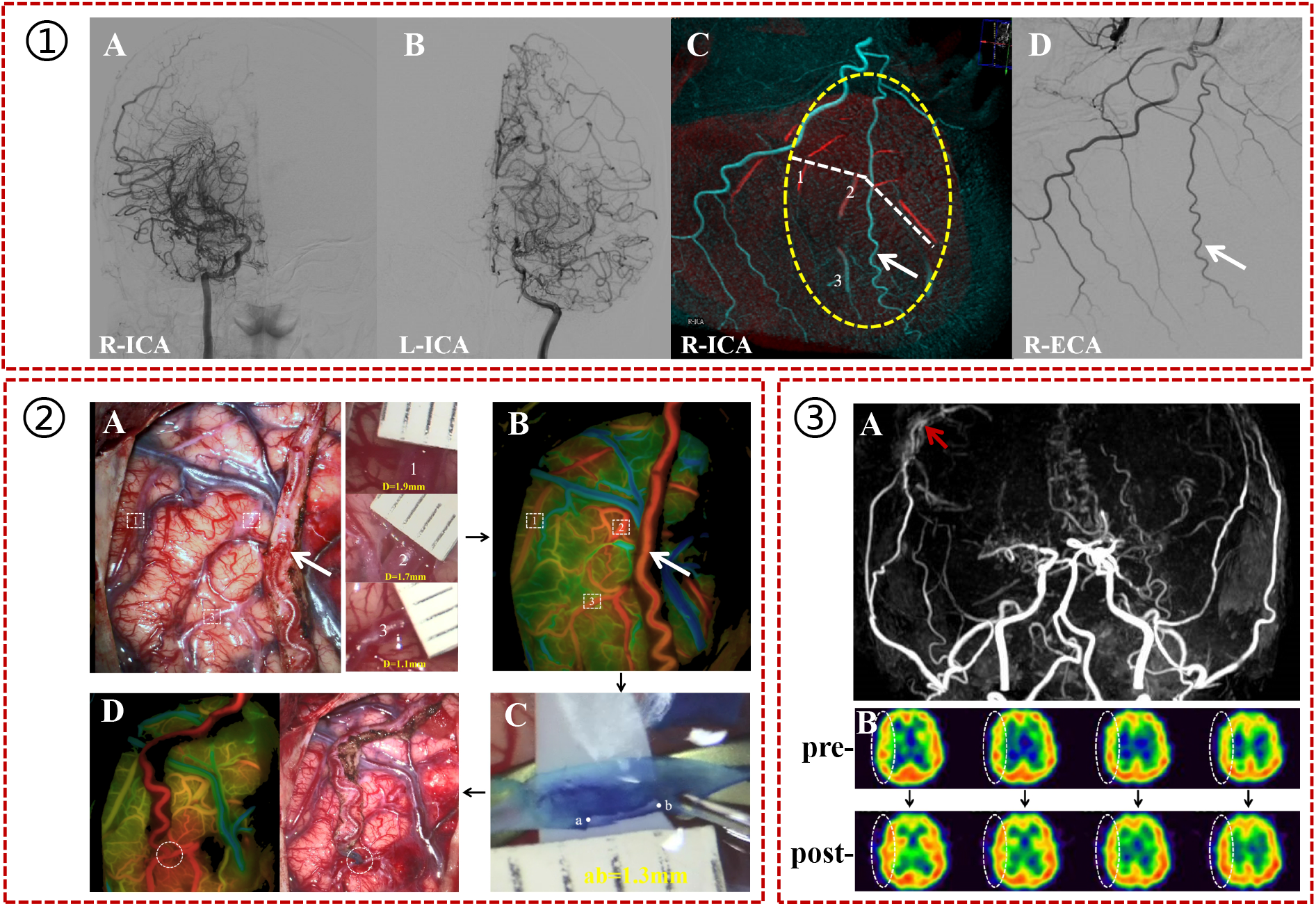
A representative case performed the flow-controlled STA-MCA anastomosis. ① (**A** and **B**) the diagnosis of MMD was established by DSA imaging. (**C**) Using 3D DSA-MR fusion imaging, the hemodynamic sources of the PSCAs in the operating field (yellow dashed oval) were clearly identified from the right MCA. Red indicates the blood on the 3D DSA images and cyan represents the blood on the MRA images. (**D**) the DSA showed two branches of STA, the thinner one (white arrow) was selected as the donor artery. **②** (**A**) the STA branch was kept patency when dissecting it from the scalp and during the craniotomy. The diameters of all potential recipient PSCAs (Number 1, 2 and 3) were 1.9 mm, 1.7 mm and 1.1 mm, respectively. (**B**) a flow 800 indocyanine green (ICG) video angiography was performed to observe the velocity difference between the donor artery and all potential recipient PSCAs. (**C**) Because the PSCA labeled No.3 had a very close flow velocity to the donor artery and only 1.1 mm in diameter, it was ultimately selected as the recipient artery and the length of the arteriotomy was designed to be 1.3 mm. (**D**) Post-anastomosis flow-800 analysis showed the patency of the anastomosis and increased cerebral perfusion around it. ③ (**A**) Postoperative MRA 2 days after surgery demonstrated the high signal intensity of the donor STA branch (red arrow). (**B**) 99mTc-ECD-SPECT 1 day after surgery revealed an overall rather than focal increase in CBF at the surgical hemisphere compared to the preoperative CBF (white dashed ovals).

## Discussion

In this study, we proposed, for the first time, a new concept of surgical revascularization which named as “flow-controlled” STA-MCA anastomosis for adult patients with MMD during performing direct anastomoses on recipient arteries with anterograde hemodynamic sources from the MCA (M-PSCAs). This “flow-controlled” anastomosis can reduce the incidence of postoperative CHP for those high-risk patients.

### Flow-controlled STA-MCA anastomosis and postoperative CHP

According to our previous study, direct anastomoses of M-PSCAs had a high risk of postoperative CHP during STA-MCA anastomosis in adult patients with MMD.^23^ The M-PSCAs were defined as PSCAs with anterograde blood flow that came from the ICA via the stenotic MCA or moyamoya vessels to the cortical arteries. Thus, the M-PSCAs represents a less efficiently hemodynamic blood flow distribution system when selected as recipient arteries.^23^ The poor vascular network related to the M-PSCAs makes the pathological vessels unable to control the suddenly and maximumly increased CBF after STA-MCA anastomosis, CHP develops in consequence.^16,19,21^ According to the long term follow up data of combined bypass for adult MMD in our center, the DSA images showed direct revascularization performed on recipient PSCAs with hemodynamic flow from the MCA provided much less contribution to postoperative flow augmentation than indirect revascularization regardless of using the “flow-controlled” or not. Thus we preferred to sacrifice part of the bypass flow in this cohort patients with a high risk of postoperative CHP. On the other hand, to ensure a success direct anastomsis, we guaranteed the diameters of the selected recipient arteries to be about 1mm. As a result, in the flow-controlled group of this study, the rate of bypass graft patency during the operation and 6 month follow up was 100% and 96.2%, respectively. The symptoms of all the ischemic MMD patients improved immediately after surgery. Although flow-controlled, improved cerebral perfusion was observed postoperatively during 6 month follow up in this cohort patients.

In the present study, the recipient arteries in less than half of the hemispheres (48.9%, 89/182) had blood flow from the MCA. Of these 89 hemispheres, the earlier 36 hemispheres were performed STA-MCA anastomoses using the traditional maximized flow augmentation concept. However, there were amazingly high incidences of focal (52.8%) and symptomatic (25%) postoperative CHP observed during their surgical procedures. Consequently, the following 53 hemispheres were performed the specifically designed flow-controlled STA-MCA anastomoses. As the result demonstrated, the incidences of focal and symptomatic postoperative CHP were significantly decreased after using the flow-controlled concept. Furthermore, the clinical presentations of symptomatic CHP in the flow-controlled group were much milder than those in the non-flow-controlled group. Multivariate analysis revealed that using the flow-controlled concept or not was significantly associated with the development of focal and symptomatic CHP. Together, these findings suggest that the flow-controlled method can significantly decrease the incidence of postoperative CHP and help to prevent the occurrence of symptomatic CHP. Our goal is to control the extent of increased intracranial blood flow for this kind of specific patients, rather than to maximize it indiscriminately.

### Basic principles of the flow-controlled STA-MCA anastomosis

As the vasculature is a dynamic and pulsatile system, the post-anastomosis hemodynamic changes may not be completely consistent with that seen in a rigid tube as the Hagen-Poiseuille equation described.^8^ Nevertheless, the correlation among the blood flow, lumen diameter, flow velocity and other parameters is still valid.^13^ In our traditional understanding of STA-MCA anastomosis, in order to improve intracranial ischemia and hypoperfusion, techniques that aimed to maximumly realize the blood flow augmentation were adopted, such as mild dilation of the STA with pressurized saline containing heparin, a arteriotomy (oblique transection of the distal STA at a 60° angle as well as a longitudinal incision in the arterial wall) to increase the area of the anastomosis site, shorten the length of the STA, selecting the stronger STA branch (if possible) as the donor artery and the cortical artery with the largest size (if possible) as the recipient artery, etc. These techniques were in accordance with the procedures introduced in the previous studies.^2,22,24^

The concept of the flow-controlled STA-MCA anastomosis is basically contrary to that of the maximized flow augmentation STA-MCA anastomosis with the aim of reducing the hemodynamic difference between the STA and recipient M-PSCAs, however, it simultaneously ensures positive blood flow to the brain. Several basic principles of the “flow-controlled” STA-MCA anastomosis should be noted based on the Poiseuille equation:

1. Controlling the “L” in the equation: First, ensuring the length of the final donor STA > 10 cm, which is similar to one reported study;^12^ As we know, a linear relationship exists between the vessel length and the resistance, which is often exploited during bypass surgery to regulate the vascular resistance, thereby adjusting the flow into the recipient vessel through the graft.^16^ Second, selecting the thinner branch of the STA as the donor artery, but ensuring its diameter > 1mm, if possible. Most of time it is easy to achieve because the mean diameter of the STA was 1.3mm.^21^
2. Controlling the “Δp” in the equation: Normally, there are several PSCAs suitable for anastomosis and the mean diameter of the PSCAs is 1.4 mm (0.8mm-1.6mm), which makes the choosing of recipient artery a difficult and confusing problem for neurosurgeons.^1,10,12^ Our “flow-controlled” method suggests selecting one with its diameter about or near to 1mm as the recipient artery for avoiding postoperative poor revascularization. In addition, we follow a principle of matching selection of donor and recipient vessels when deciding the recipient artery.^9^
3. Controlling the “r” in the equation: According to the Hagen-Poiseuille equation, a change in radius alters resistance and it therefore has the biggest impact on flow. Due to lack of reference standards, we designed the length of the arteriotomy less than 1.2-fold the diameter of the recipient artery. This ensures positive blood flow from STA to the brain but not excessive.

### Limitations

Our study is limited by its retrospective nature. Moreover, it is a single-center study which creates regional bias of the sample which could be an issue. As FLOW 800 analysis allows only detection of procedure-related hemodynamic changes within the microcirculation and macrocirculation but not assessment of quantitative perfusion or flow,^17^ precise flow evaluation was absent in this study. Large sample size studies with accurate flow measurement are needed in the future.

## Conclusions

In this study, we suggested a new concept of STA-MCA anastomosis for adult MMD, which named as flow-controlled anastomosis. As a complementary method to the traditional direct bypass procedure, the flow-controlled anastomosis ensures a “controlled”, rather than a “maximized” augmentation of blood flow through reducing the hemodynamic difference between the STA and recipient M-PSCAs, and consequently decreases the incidence of postoperative CHP in adult patients with MMD.

## Data Availability

All data produced in the present work are contained in the manuscript

## Acknowledgments

This study was partly supported by two projects of the National Natural Science Foundation of China (81671157 and 8217052644).

